# First Use of Phage Therapy in Canada for the Treatment of a Life-Threatening, Multidrug-Resistant *Staphylococcus epidermidis* Periprosthetic Joint Infection: An N-of-1 Trial

**DOI:** 10.1101/2025.02.21.25321539

**Authors:** Melissa T. Cammuso, Bradley W. M. Cook, D. William Cameron, Stephen Ryan, Gina A. Suh, Marisa A. Azad

## Abstract

We describe the first use of phage therapy in Canada for the treatment of a life-threatening periprosthetic joint infection (PJI), with successful outcome. PJI is a devastating complication of joint replacement surgery, with high morbidity and mortality. Our patient presented with early sepsis from a chronic recalcitrant multidrug-resistant *Staphylococcus epidermidis* hip PJI which had repeatedly failed standard therapy. She had previously undergone 10 operations of the right hip, and only three weeks after completing a prolonged course of daptomycin following her most recent hip revision, she developed a draining sinus tract. Given the high burden of disease, inability to achieve surgical source control, and lack of antibiotic treatment options for long-term suppressive therapy, bacteriophage (phage) therapy was pursued. The patient underwent irrigation and debridement with complex flap reconstruction: intraoperative tissue cultures again yielded MDR *S. epidermidis*. We developed a novel phage protocol for this patient, with twice daily, intra-articular and intravenous (7 × 10^9^ PFU/dose) phage delivery over a planned 14-day course. Complete healing of the wound with cessation of drainage occurred within one month after treatment. A marked improvement in right hip pain and mobility occurred within three months after treatment. Twelve months following phage treatment, there is normalization of serum inflammatory markers with diminished pain, increased mobility, and no recurrent surgery. Our patient continues to improve and is currently living independently at home, with sustained clinical control of infection.

## INTRODUCTION

A female patient in their 70s presented to our periprosthetic joint infection (PJI) clinic with a one-week history of subjective fever and a draining abscess overlying the right hip (**Figure 1**). Her past medical history was significant for drug reaction with eosinophilia and systemic symptoms (DRESS; β-lactam and glycopeptide antibiotic classes were to be avoided). Her orthopedic surgery history was extensive, including 10 operations of the right hip, with development of a recalcitrant multidrug resistant (MDR) *Staphylococcus epidermidis* right hip PJI (antibiotic susceptibilities shown in **Table 1**). She had failed treatment with multiple courses of antibiotics, including chronic suppression with doxycycline, and – just prior to presentation – a 12-week course of daptomycin with concurrent revision of the right total hip arthroplasty and acetabular reconstruction. While on daptomycin, she developed peripheral neuropathy and myalgias.

**Table 1.** *Staphylococcus epidermidis* isolate extended antibiotic susceptibility profile.

| Antibiotic | *Staphylococcus epidermidis* Minimum Inhibitory Concentration (MIC) or Zone of Inhibition | Interpretation |
| --- | --- | --- |
| Oxacillin | 2 mg/L | Resistant |
| Clindamycin | >4 mg/L | Resistant |
| Ciprofloxacin | 7 mm | Resistant |
| Rifampin | ≤0.25 mg/L | Susceptible |
| Tetracycline | 6 mm | Resistant |
| Trimethoprim/Sulfamethoxazole | 4 mg/L | Resistant |
| Vancomycin | 1 mg/L | Susceptible |
| Daptomycin | ≤0.5 mg/L | Susceptible |
| Linezolid | ≤2 mg/L | Susceptible |
| Ceftaroline | 0.38 mg/L | Unknown* |
| Ceftobiprole | 0.75 mg/L | Unknown* |
| Fosfomycin | 16 mg/L | Unknown* |

\*Note that no Clinical and Laboratory Standards Institute (CLSI) minimum inhibitory concentration (MIC) breakpoints currently exist.

**Figure 1.**
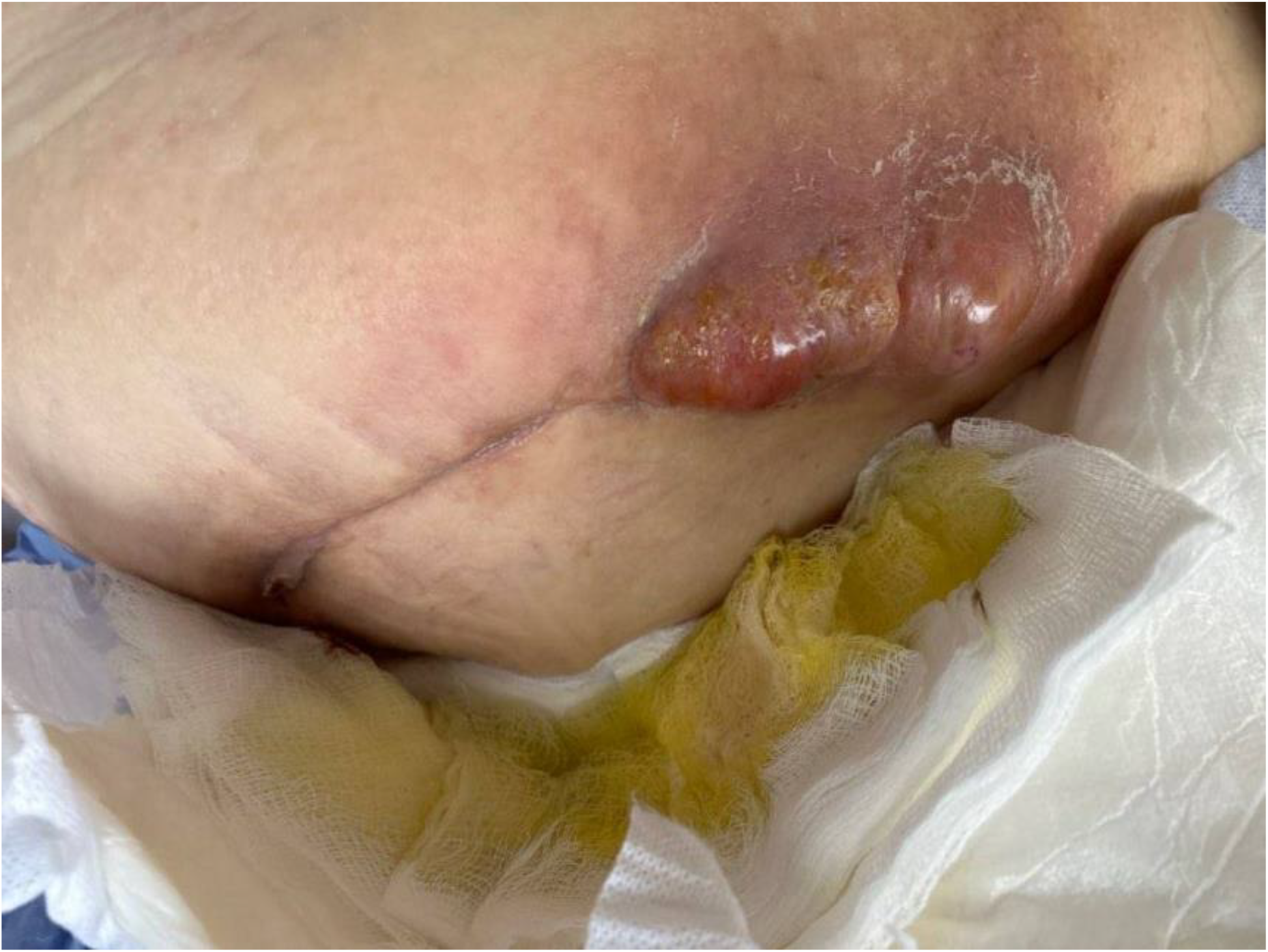
Development of a *S. epidermidis* abscess with a draining sinus tract overlying the right hip three weeks after cessation of a 12-week course of daptomycin.

Only three weeks after completion of daptomycin, she developed increasing fatigue, right hip pain, subjective fever, rigors, and a draining sinus tract overlying the right hip posterior incision site. C-reactive protein (CRP) and erythrocyte sedimentation rate (ESR) were 85 mg/L and 67 mm/h, respectively. Creatinine kinase was within normal limits. The patient was afebrile but appeared unwell and lethargic. Copious purulent fluid was draining from the right hip sinus tract. A pelvic X-ray and computed tomography (CT) of the right hip are shown in **Figures 2** and **3**, respectively. She was promptly readmitted. A goals of care discussion was held: given that surgical source control with complete implant removal (i.e., a right hemipelvectomy) could not be safely achieved, and viable antibiotic treatment options for long-term suppressive therapy were not available, bacteriophage (phage) therapy was pursued. The patient underwent irrigation and debridement with complex flap reconstruction and was restarted on daptomycin with the addition of oral ciprofloxacin empirically, pending microbiology. All four intraoperative tissue cultures yielded MDR *S. epidermidi*s. Antimicrobial therapy was transitioned to daptomycin and oral rifampin. She re-developed myalgias and peripheral neuropathy on daptomycin, even at reduced dosing.

**Figure 2.**
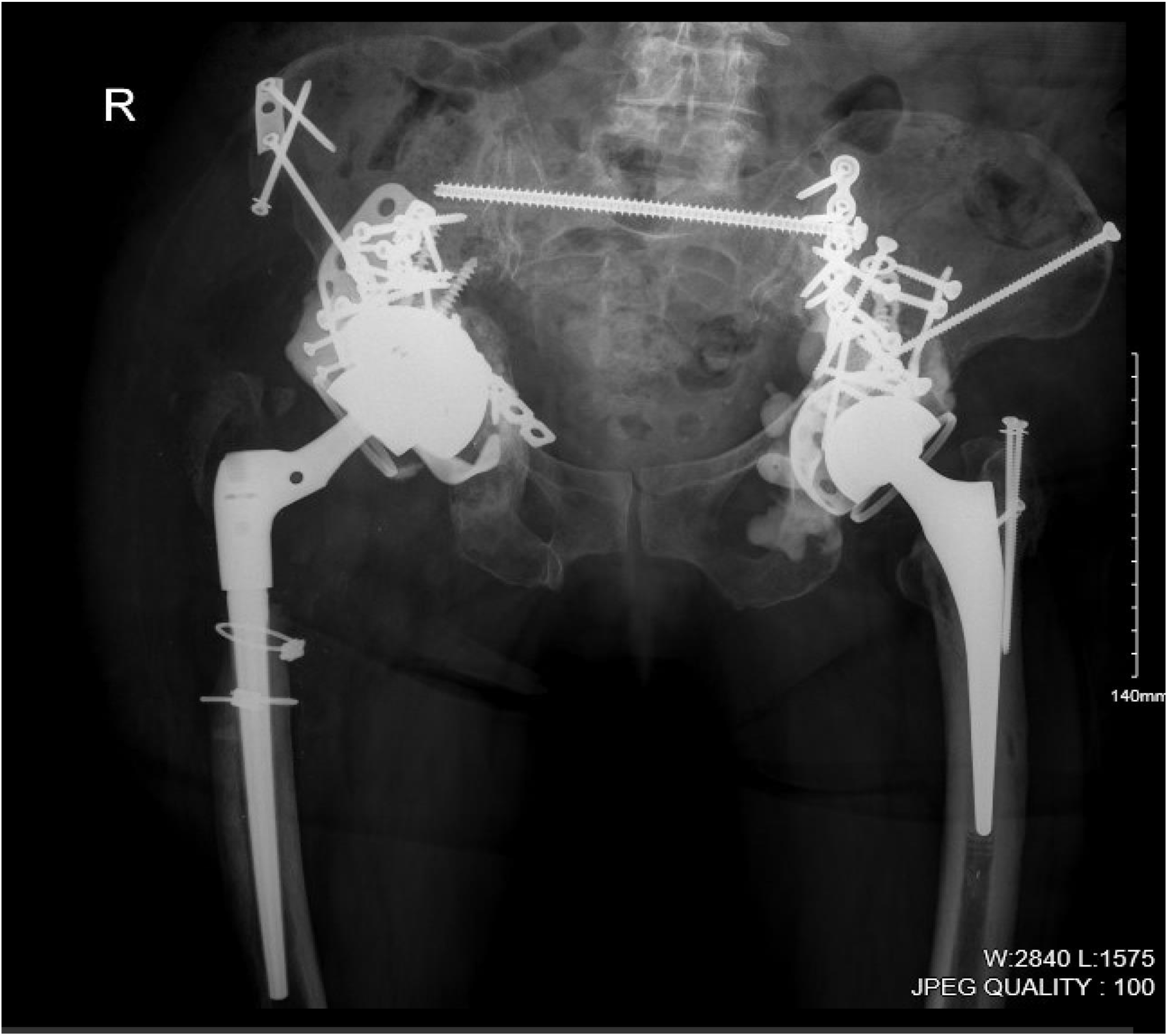
Pelvic X-ray highlighting the burden of *S. epidermidis* infected hardware involvement of the right hip and hemipelvis.

**Figure 3.**
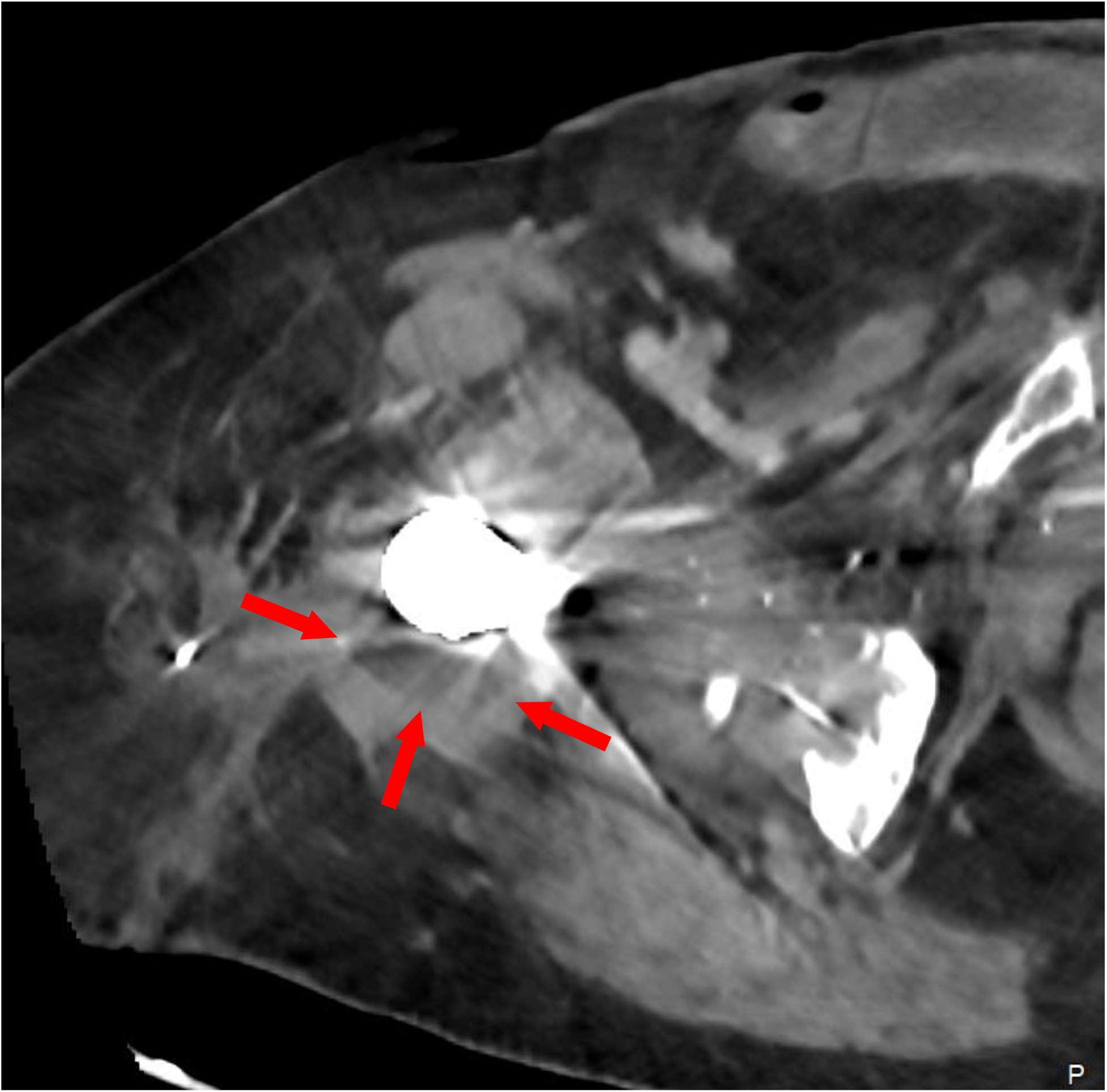
Axial CT image of the right hip demonstrating a stable periprosthetic fluid collection (red arrows) post percutaneous drainage catheter insertion (catheter not imaged).

### Phage Therapy

In collaboration with Cytophage Technologies, Ltd., (Winnipeg, Canada), we developed a novel phage protocol for this patient. Regulatory, research ethics board, and institutional approvals from Health Canada and the Ottawa Hospital Research Institute (OHRI) were obtained (OHRI Protocol#: 20240008-01H). Preclinical *in vitro* testing against the *S. epidermidis* isolate led to the identification and characterization of the therapeutic potential of a single, lytic phage. The phage was not cultivated using the clinical isolate, as whole-genome sequencing identified two intact prophages which could contaminate final phage preparations. The phage was propagated within an avirulent *Staphylococcus succinus* host. Resultant phage lysates were concentrated and purified. The therapeutic quality monophage preparation was profiled using rigorous testing by Cytophage and a third-party, thus ensuring final product potency (7 × 10^9^ PFU/mL), purity, fidelity, and safety. These tests included United States Pharmacopeia (USP) standards (sterility, endotoxin and physical chemistry) and final product identity measures (electron microscopy, PCR, and sequencing), in adherence to compassionate-use recommendations (1).

## RESULTS & DISCUSSION

### Results

The patient was admitted, underwent ultrasound-guided percutaneous drainage catheter insertion into a right hip fluid collection, and was administered phage therapy, twice daily, both intra-articularly and intravenously (7 × 10^9^ PFU/dose). She received concomitant daptomycin and oral rifampin. The catheter became malpositioned on day seven of treatment which was then removed to prevent the introduction of additional microorganisms into the joint space (intra-articular phage administration was discontinued). Intravenous phage therapy was continued for an additional seven days, completing the planned 14-day treatment course. After phage administration, the patient developed initial and immediate mild hypotension, followed by hypertension, low-grade fever, chest pain, rigors, and wheezing, lasting approximately 30 minutes from treatment on days one to five. These reactions were more marked following intra-articular phage delivery which followed intravenous administration, peaking after the second dose on day one, then subsiding with each subsequent dose, with resolution by day five. Liver enzymes (alanine transaminase [ALT], aspartate aminotransferase [AST], alkaline phosphatase [ALP], and gamma-glutamyl transpeptidase [GGT]) increased and peaked in the first week of treatment (baseline and peak values: AST 30 to 99 U/L; ALT 29 to 108 U/L; ALP 124 to 223 U/L; GGT 61 to 105 U/L). Liver enzymes began to fall at week two of treatment and returned to baseline by one month after treatment. Baseline inflammatory biomarkers, ESR and CRP, were 51 mm/hr and 28 mg/L, respectively. CRP peaked on day 3 of treatment (93 mg/L) and decreased to 43 mg/L at one-month follow up. ESR remained overall unchanged throughout treatment.

The patient was monitored in PJI clinic. Complete healing of the wound with cessation of drainage occurred within one month after treatment. A marked improvement in right hip pain and mobility occurred within three months after treatment. Daptomycin was transitioned to oral linezolid three months after treatment and rifampin was discontinued after seven months. An overall decrease in serum CRP and ESR occurred with normalization 12 months after treatment. **Table 2** shows an overall trend in inflammatory markers. There is sustained improvement in right hip pain and mobility, and the patient has moved from family-assisted living to independent living in her own home. Given how high risk this patient is for relapse of infection (with significant associated mortality and morbidity), a decision was made to continue linezolid, with close monitoring for toxicity. Should this occur, three-times weekly linezolid will be considered, with eventual cessation of therapy, assuming ongoing clinical improvement and/or stability, near the one-year mark. A repeat hip aspirate will be considered then.

**Table 2.** Overall trend of inflammatory markers during and after phage treatment.

|  | Baseline | Week 1 of Treatment | Week 2 of Treatment | Month 6 Follow-up | Month 12 Follow-up |
| --- | --- | --- | --- | --- | --- |
| CRP (mg/L) | 28 | 64 | 34 | 23 | 2.4 |
| ESR (mm/hr) | 51 | 55 | 46 | 43 | 5 |

## Discussion

PJI is a catastrophic disease which complicates 1-2% of primary joint replacements (2). In Canada, PJI is one of the leading causes of revision operations in hip and knee replacements and poses a large burden of disease (3). Early recognition, accurate microbiologic diagnosis, and prompt surgical and medical treatment of PJI are critical to preventing recalcitrant infection, which is not only incurable, but associated with significant morbidity and mortality (4). Frequently, chronic PJI requires long-term antibiotic therapy and multiple surgical interventions. In severe cases, resection arthroplasty or amputation may be needed. There is an unmet need for novel therapeutic approaches.

Phage therapy is underexplored for PJI. Phage are viruses which are ubiquitous in nature and specifically infect bacteria. In specific cases, phage is an attractive adjunctive therapeutic option for targeted antimicrobial therapy, and for combating antimicrobial drug resistance. Overall, promising outcomes with phage therapy as a treatment for PJI have been summarized in recent reviews (5, 6). In all cases, phage therapy was delivered to patients that repeatedly failed standard treatment options. Phage treatment protocols were variable with respect to preparations (monophage or cocktails), dosing (single or sequential; concentration), frequency and duration, method of delivery (e.g., intra-articular, topical, and/or intravenous), concurrent surgical intervention, co-administration of antibiotic(s), and follow-up. In a systematic review by Yang et al., 16 clinical studies were screened (one prospective controlled clinical trial and 15 case reports), in total including 42 patients with hip and/or knee PJI who received phage therapy. Following phage therapy, two cases demonstrated relapse of infection, while others demonstrated improved outcomes (5). In a systematic review by Suh et al., the outcomes of 33 bone and joint infection cases treated with phage therapy since 2010 were highlighted. Of these cases, 87% achieved favorable outcomes (microbiologic or clinical success), with only rare mild adverse events (6). Overall, despite having non-standardized protocols, phage therapy appears to be a potential adjunctive modality to treat orthopedic infections, including PJI.

Phage therapy is generally considered safe, although rare and minor adverse events have been reported such as local reactions at the treatment site (erythema, pain), transient pruritis, shortness of breath and wheezing, fevers and chills, flushing, and hypotension (7, 8, 9). Our patient demonstrated an interesting tempo of reactions during phage treatment which have not, to our knowledge, been reported, including initial and transient hypotension, followed by hypertension, low-grade fever, chest pain, rigors, and wheezing during early treatment. These were worse following initial intra-articular phage delivery (which followed intravenous injections) and resolved by day five of treatment. Little is known about the immunobiology of these reactions, although data may point towards the release of endotoxins (in cases of Gram-negative bacterial infection) or other bacterial components (e.g., DNA, lipoteichoic acid, membrane-embedded proteins, and secreted exotoxins) as stimulators of inflammatory cytokine responses (10).

We report the first use of phage therapy in Canada for the treatment of a life-threatening hip PJI, with successful outcome of wound healing, diminished pain, increased mobility, and normalization of inflammatory biomarkers, without recurrent surgery. Our patient continues to improve and is currently living independently at home, with sustained clinical control of infection. Several barriers to the wide-spread use of phage therapy exist, including regulatory hurdles and a lack of standardization and high-quality evidence to better understand which PJI patients would benefit most from this therapy. Despite these challenges, phage therapy appears to be a promising potential adjunct for recalcitrant PJI where standard medical and surgical management has repeatedly failed.

## Data Availability

All data produced in the present work are contained in the manuscript.

## ACKNOWLEDGEMENTS

The authors thank the Cytophage Technologies Inc. laboratory team whose efforts were critical for the experimental treatment (Marielou Tamayo, Melissa Plett, Stephanie Lau, Tia Arnaud, Dr. Henrik Almblad, Nicolas Fournier, Karen LoVetri, Dr. Tasia Lightly, Dr. Yuen Ming Chung, Riya Roy, Natasha Theriault, and Dr. Steven Theriault). The authors also thank Dr. Jessica Sacher for fruitful discussion. We also wish to thank Dr. John Bell, Dr. Jennifer Quizi, Dr. Mathieu J.F. Crupi and the Biotherapeutics Manufacturing Centre (Jessica Hentschel), as well as our microbiology colleagues at Eastern Ontario Regional Laboratory Association.

